# Effect of Kangaroo Mother Care during the first 72 hours of life on early growth and breastfeeding in normal birth weight newborns: Protocol for a Randomised Controlled Trial

**DOI:** 10.64898/2026.02.23.26346760

**Authors:** Aarti Kumar, Malvika Mishra, Madhuri Tiwari, Vinay Pratap Singh, Rupak Mukhopadhyay, Anushka Srivastava, Mala Kumar, Anjoo Agarwal, Shakal Narayan Singh, Shalini Tripathi, Mohd. Salman Khan, Pradeep Kumar, Rashmi Kumar, Ashok Kumar, Gary Darmstadt, Vishwajeet Kumar

## Abstract

**Introduction:** Skin-to-skin contact during the first hour of birth is recommended for healthy newborn infants and their mothers and improves early stabilisation and breastfeeding outcomes. Kangaroo Mother Care (KMC), involving prolonged skin-to-skin contact (SSC) and exclusive breastfeeding, provides an optimal physiological transition from intrauterine to extrauterine life for preterm and low birth weight infants, improving survival by 32% and conferring multiple clinical and neurodevelopmental benefits. Extending the duration of SSC contact beyond the first hour may similarly confer underexplored benefits to normal birth weight infants, including improved stabilisation, breastfeeding, growth, and maternal-infant bonding. The study aims to evaluate the effect of KMC during the first 72 hours on early weight loss, weight gain velocity and breastfeeding quality in normal birthweight infants.

**Methods and Analysis:** This multicentre, individually randomised, controlled, open-label superiority trial will enrol 516 healthy singleton neonates with birth weight ≥2500 grams and their mothers with uncomplicated vaginal deliveries from 3 public health facilities in Uttar Pradesh, India. Dyads will be randomised (1:1) and stratified by site to either intervention or control groups. The intervention involves prolonged KMC (≥8 hours of daily SSC with exclusive breastfeeding in the KMC position) during the initial 3 days after birth, with a recommendation to continue KMC at home throughout the newborn period. Both intervention and control groups will receive a common minimum care package, including breastfeeding initiation through uninterrupted SSC in the first hour, essential newborn care counselling, vaccinations and other standard facility care. The primary outcomes are: 1) mean percentage weight loss at 48 hours; 2) weight gain velocity up to 28 days; and 3) the proportion of dyads with moderate-to-poor quality breastfeeding scores (BBAT <7) at age 7 completed days. Secondary outcomes include exclusive breastfeeding rates, maternal breastfeeding experience, incidence of possible serious bacterial infection, maternal depression, and maternal-infant bonding. Data will be collected electronically using standardised tools with quality control measures. Primary outcomes will be analysed using Linear Mixed-Effects Models (continuous) and Mixed-Effects Logistic Regression (binary) on an Intention-to-Treat basis, adjusting for study site, parity, infant sex, and baseline birth weight. A p-value <0.05 will be considered statistically significant.

**Ethics and Dissemination:** The study is approved by the institutional ethics committees of the Community Empowerment Lab and King George’s Medical University. Written informed consent will be obtained from participating mothers. All findings will be disseminated regardless of the outcome, through publication in peer-reviewed journals, presentations at international conferences, and policy briefings to local health authorities. Data will be deposited in an open-access repository to promote data sharing and transparency. The results are intended to inform national and international guidelines on essential newborn care for the global population of healthy, term infants.

**Strengths and limitations of the study:**

- This study fills an important evidence gap, being the first multi-centre randomised controlled trial to evaluate prolonged KMC benefits for normal birthweight infants, a group that has largely been excluded from prior KMC research.
- The study will conduct a rigorous and objective assessment of a range of important newborn and maternal outcomes, including physical growth, quality of breastfeeding and exclusive breastfeeding, maternal mental health and mother-infant bonding, providing a holistic understanding of KMC’s effects on the newborn-mother dyad.
- The visible nature of KMC makes blinding participants and implementers impossible, however, extensive measures have been undertaken to minimise bias. Being the first study of its kind, it excludes Caesarean section and complicated births to simplify the intervention, analysis and subsequent interpretation.

**Administrative information:** *Title:* Effect of Kangaroo Mother Care during the first 72 hours of life on early growth and breastfeeding in normal birth weight newborns: Protocol for a Randomised Controlled Trial

*Trial registration:* This trial has been registered with the Clinical Trials Registry of India CTRI/2024/01/062057 (date: 30/01/2024) and International Standard Randomised Controlled Trial Number ISRCTN14346778 (date: 19/12/2024).

*Protocol date and version:* September 10, 2024; CEL/KMC_NormalBW/V3.1. The full study protocol and statistical analysis plan will be available as supplementary material accompanying the published manuscript. The protocol has also been uploaded to the CTRI registry.

*Funding & Role of sponsor:* This work was supported by Indian Council of Medical Research (Project number: IIRP-2023-7329). The sponsor has no role in the design, conduct, analysis and reporting of the trial, but provides study oversight through a scientific review committee.

*Sponsor contact information:* Indian Council of Medical Research (ICMR), New Delhi – 110029, India

## INTRODUCTION

The transition from a protected foetal environment to independent newborn life is a period of profound physiological change. The warm, fluid-filled sanctuary of the womb provides the foetus with vital oxygen, a thermally stable environment, continuous nutrition through placental glucose, and immunological protection. At birth, a sudden exposure to an unregulated, cold, dry, pathogen-loaded environment without the vital support of the placenta requires rapid respiratory, thermoregulatory, metabolic, cardiovascular, skin-barrier, and immune-system adaptations.^1,2^

Although early vital transition needs to occur within the first 2-6 hours of life in healthy term neonates, critical adaptive processes stabilise over 24-72 hours.^1,3^ Successful and expeditious transition depends on both intrinsic factors, such as foetoplacental health and gestational age at birth, and extrinsic factors, including mode of delivery, newborn care and exposure to environmental stressors.^4–6^ The energy requirement for adaptation varies with these factors and is met through internal energy stores supplemented by external nutrition, e.g. via breastfeeding.^7^ Sub-optimal care practices like separation from the mother and environmental stressors can exacerbate risks to the newborn, contributing to cycles of instability that may delay or even prevent successful adaptation.^8,9^ Further, the first 48-96 hours are also a crucial period for establishing successful breastfeeding, a practice that plays a critical and multifaceted role in supporting newborn adaptation to extrauterine life by kickstarting their immune defence, stabilising their metabolism, and providing thermal and emotional regulation.^10,11^ Frequent maternal-newborn separation, poor positioning and latching contribute to initial feeding challenges with inadequate milk intake, hypoglycaemia and frequent crying fuelling maternal perceptions of insufficient milk production.^12–15^ This often results in pre-lacteal and supplemental feeding during the initial days, undermining breastfeeding success, disrupting adaptation and increasing morbidity risk.^16,17^

Uninterrupted skin-to-skin contact (SSC) during the first hour of life has been shown to support breastfeeding initiation and continuity and to improve early cardiorespiratory and metabolic stabilisation, and is recommended by the World Health Organization (WHO) for all healthy term newborns.^18^ This practice is often clinically differentiated from Kangaroo Mother Care (KMC), recommended by the WHO for low-birthweight and preterm infants, with more than 8 hours of daily SSC with exclusive breast milk feeding over several days and weeks.^19^ In this subgroup of infants, KMC improves survival by 32%, and provides other key benefits, including improved thermoregulation, physiological stabilisation, maternal-infant bonding, reduced postpartum depression, and neurodevelopmental benefits that are sustained at 20-year follow-up.

Strong evidence of the benefits of KMC in low-birthweight and preterm infants supports the biological plausibility that continuing SSC beyond the first hour may confer similar benefits to normal birthweight newborns and their mothers, but remains underexplored. Limited evidence suggests that KMC practised during hospital stay may be beneficial in preventing hypothermia at discharge,^20^ and for managing hyperbilirubinemia.^21^ The KMC position is known to elicit primitive neonatal reflexes in healthy term newborns, facilitating breastfeeding as an innate behaviour and improving latching, thereby supporting increased milk intake. ^22,23^ SSC in full-term neonates practised throughout the neonatal period may promote exclusivity of breastfeeding and improve the mother-infant relationship.^24^

This trial aims to systematically investigate the effects of prolonged KMC with breastfeeding in the KMC position over the first 72 hours of life in healthy, normal-birthweight newborn-mother dyads on early weight loss, weight gain velocity, breastfeeding quality, and other key newborn and maternal outcomes.

## OBJECTIVES

### Primary objectives

To evaluate the effect of prolonged KMC (at least 8 hours and ideally

>20 hours of daily SSC along with exclusive breastfeeding in the KMC position) in the first 72 hours of life among normal birth weight infants on:

1. mean percentage weight loss observed during the first 2 days of birth
2. mean weight gain velocity during the newborn period i.e. the first 28 days of birth
3. proportion of mother-baby dyads with moderate-to-poor Bristol Breastfeeding Assessment Tool (BBAT) scores (<7 out of 8) at age 7 completed days

### Secondary objectives

To compare exclusive breastfeeding, mother’s breastfeeding experience and self-efficacy, possible serious bacterial infection (PSBI), maternal depression and mother-infant bonding between intervention and control groups over a 28-day follow-up period.

## METHODS: TRIAL DESIGN, PARTICIPANTS, INTERVENTIONS & OUTCOMES

### Research Hypothesis

Prolonged Kangaroo Mother Care (KMC) (≥8 hours daily SSC with exclusive breastfeeding) during the first 72 hours of life among infants weighing ≥2500 grams at birth, as compared with standard care, will lead to:

a. 25% reduction in mean percentage weight loss during the first 2 days of life,
b. 20% improvement in mean weight gain velocity during the first 28 days, and
c. 50% reduction in the proportion of mother-baby dyads with moderate-to-poor Bristol Breastfeeding Assessment Tool (BBAT) scores (<7 out of 8) at age 7 completed days.

### Study Design

This is a multicentre, pragmatic, individually randomised, controlled, open-label superiority trial comparing two parallel groups of normal birthweight infants, (weighing ≥ 2500 grams at birth) - early and prolonged KMC with breastfeeding in the KMC position for the first 72 hours after birth (intervention group) WITH standard care (control group). Both groups will receive a common minimum package of essential newborn care components. The unit of randomisation is the mother-infant dyad in a 1:1 ratio. The Consolidated Standards of Reporting Trials (CONSORT) flowchart of the study is illustrated in Figure 1.

**Figure 1.**
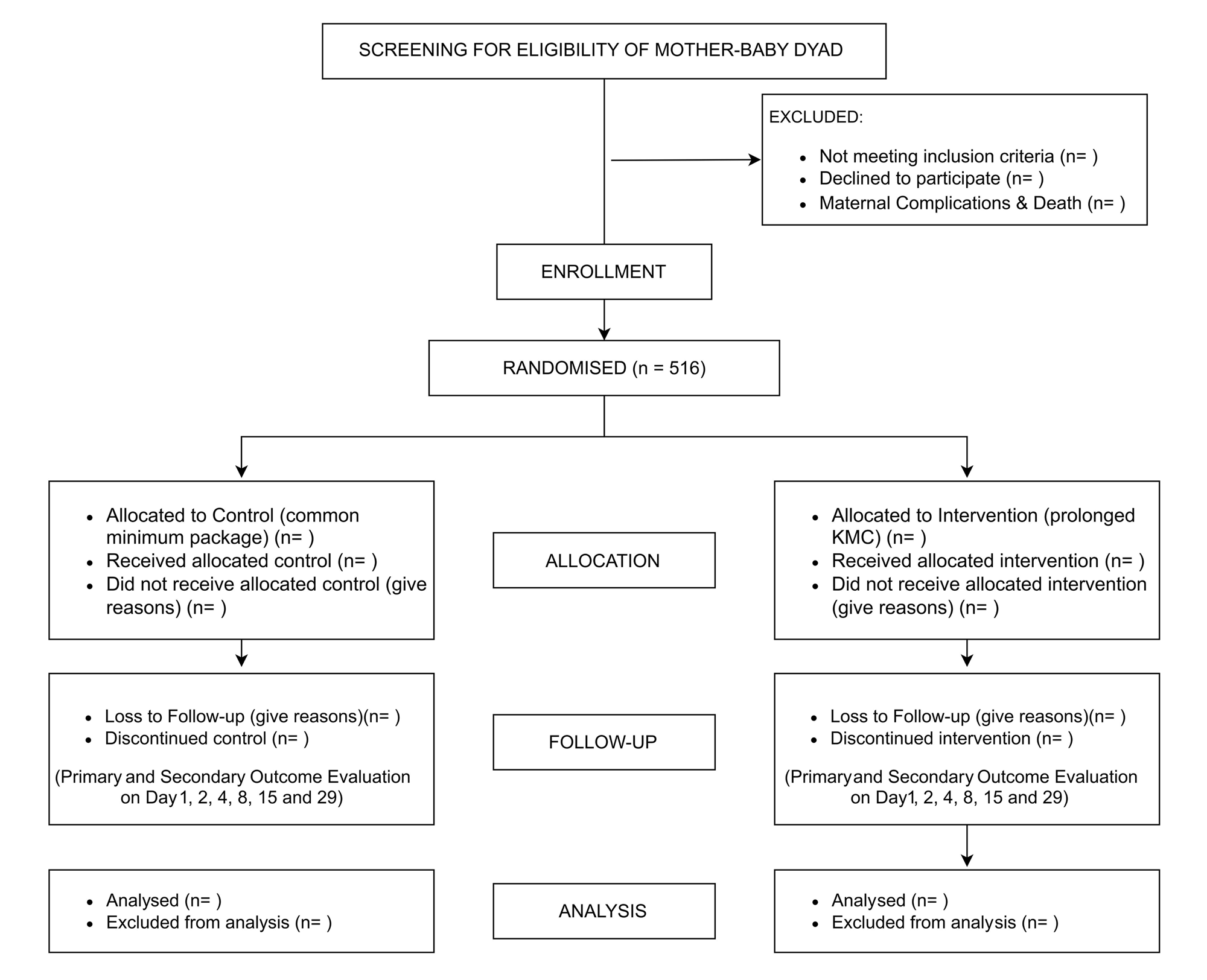
Study CONSORT Flow Diagram

### Study Setting

The multicentre study will be conducted in three public facilities in and around Lucknow, Uttar Pradesh, India – Queen Mary Hospital (a tertiary-level public facility operated by King George’s Medical University), Veerangana Avanti Bai District Women’s Hospital, and District Women’s Hospital, Barabanki. The selection of study sites is based on expected enrolment rates, feasibility of segregating the intervention and control mothers, and other pragmatic considerations.

### Patient and Public Involvement

No formal patient or public involvement is anticipated in the design, conduct or reporting of the trial. We may involve intervention mothers and families during the dissemination of study findings to the broader public.

### Study Population

The study will include healthy singleton neonates with a birth weight of 2500 grams or more, delivered vaginally and without complications at the study facility. Eligible mothers must reside within a catchment area radius of about 50 kms, agree to stay in the facility for at least 48 hours post-delivery, intend to breastfeed for the first 28 days, and must provide written informed consent within 4 hours of birth.

Exclusions: Neonates requiring immediate NICU admission after birth, enrolled in other studies or with major congenital malformations apparent at birth that could interfere with provision of the intervention (e.g. anencephaly, hydrocephaly, cleft lip/ palate, meningomyelocele, omphalocele, imperforate anus) will be excluded. Further, newborn-mother dyads where mothers die prior to screening, require special or intensive care due to complications, have multiple births, caesarean section as the mode of delivery, are HIV+ positive and planning replacement feeding, or have generalised skin conditions likely to hinder prolonged KMC (e.g. rashes, infections, etc.), will be excluded.

### Intervention

#### Prolonged KMC

The intervention will involve a minimum duration of 8 hours and will target up to 20 hours of daily SSC during the first 3 days, with exclusive breastfeeding in the KMC position (similar to WHO recommendations for low birth weight and preterm infants).^26^ During SSC, as per KMC protocols, the newborn and the mother will remain bare-chested, with the baby’s head covered by a cap, and socks for warmth, and a blanket placed over the baby to cover its back and the mother’s chest. The mother will be positioned semi-reclined at an angle of 30-45 degrees. Mothers may also provide ambulatory KMC with the help of a binder. After the initial 3-day period, mothers will be encouraged to continue KMC and breastfeeding in the KMC position to the extent they are comfortable, throughout the 28-day newborn period. Mothers are encouraged to be the primary providers of KMC, with some support from family members to increase the dose of SSC.

#### Breastfeeding in KMC position

Breastfeeding sessions will be newborn-led, implying on-demand feeding, with the newborn attempting self-latch in the KMC position. With the baby in the KMC position, the mother will be encouraged to position and support her breast, so it does not move as the baby attempts to latch on its own without interference from the mother. This approach leverages the KMC position to improve latching-on by utilising the newborn’s primitive reflexes, facilitating breastfeeding as an innate rather than learned behaviour and avoiding common mistakes mothers inadvertently make in attempting to support their babies.^22,23^

#### Role of KMC intervention support team

A team of lay workers trained in KMC support (KMC team) will provide counselling and hands-on support to mothers in practising the intervention, as described. During the initial hours after enrolment, the KMC team member will explain the benefits of KMC to intervention group mothers and teach them how to keep their babies safely in the KMC position. Once the baby is in KMC, they will wait for the baby to show hunger cues and teach the mother how to allow the baby to self-latch in the KMC position, without interfering with the process. They will also teach the mother to use a binder to secure their baby in the KMC position and practice ambulatory KMC. At discharge, they will counsel the mother and family on continuing the intervention for at least one more day, but ideally throughout the newborn period. Further, they will make a telephonic call to the mother on the day after discharge to remind her to continue the intervention, as advised.

#### Criteria for discontinuing intervention

The intervention may be discontinued if the mother or newborn develops a medical condition requiring physical separation (e.g., maternal complications requiring intensive care, newborn illness requiring admission to a neonatal intensive care unit), or upon maternal request. Mothers and babies for whom the intervention is discontinued will continue to receive standard facility care and will remain in the study for follow-up outcome assessments per protocol.

#### Common minimum package for both the study groups

Both the study groups will receive standard care as per facility norms from facility health providers, including immediate newborn care with initiation of breastfeeding through uninterrupted SSC during the first hour, weighing and general assessment, routine counselling, vaccinations, physician rounds, documentation of case records, etc. The SESS team will strengthen core care processes at the study facilities intended for both groups of study participants and will be present during the birth to supervise breastfeeding initiation and accurate birthweight measurement. Given sub-optimal counselling by facility health providers, the SESS team will provide a standardised counselling package on essential newborn care to mothers in both study groups to ensure a minimum standard of counselling for all mothers. This counselling package includes thermal care (without mentioning SSC, teaching optimal wrapping of the newborn and delayed bathing), breastfeeding (hunger cues, characteristics of good latch, exclusive breastfeeding), hygiene (handwashing, cord care), danger sign recognition and appropriate care-seeking. Optimal ambient temperature and privacy in the labour room and wards will be ensured. All mothers will receive a “baby kit” containing a front-open shirt, cap, mittens, socks, blanket, diapers, handwash and hand sanitiser.

### Study Outcomes and Measurement

The primary outcome measures include percentage weight change over the first 48 hours after birth, weight gain velocity during the first 28 days of life, and quality of breastfeeding at 7 days of age, as assessed using the Bristol Breastfeeding Assessment Tool (BBAT) (as defined in Table 1). Key secondary outcome measures include exclusive breastfeeding, breastfeeding experience, possible serious bacterial infection, maternal depression, mother-to-infant bonding, and breastfeeding self-efficacy (as defined in Table 2).

**Table 1.** Primary Outcome Measures.

| Name | Definition |
| --- | --- |
| **Outcome 1.1:** Percentage weight change over the first 48 hours after birth. | **Description:** Percentage weight change over the first 48 hours after birth is defined as:<br>$\frac{\text{Weight taken at } (48 \pm 6) \text{ hours} - \text{Birth weight}}{\text{Birth weight}} \times 100$<br>**Measurement:** Weighing at birth will be done by facility nurses under direct observation of the SESS team, and at 48±6 hours after birth by the IOA team, using a calibrated Seca 334 digital weighing scale accurate to ±5 grams. Measurements will be taken by trained personnel following a uniform anthropometry protocol, ensuring calibration of the scales every 14 days. |
| **Outcome 1.2:** Weight gain velocity during the 28-day newborn period | **Description:** Weight gain velocity (g/kg/day) will be calculated as:<br>$\frac{\text{Weight at day } n - \text{Birth weight}}{\text{Birth weight}} \times \frac{1}{n} \times 1000$<br>where all weight measures are in grams, and n is the infant's completed age (in days) at the last follow-up, between 28-32 days (this range takes into account slight variations in day of follow-up due to field conditions, availability of the family, etc.)<br>**Measurement:** Weight at the last follow-up visit will be measured by trained IOA assessors using a calibrated Seca 334 digital weighing scale, ensuring consistency in measurement procedures. |
| **Outcome 1.3:** Moderate-to-poor Bristol Breastfeeding Assessment Tool (BBAT) score (<7 out of 8) at 7 completed days of age | **Description:** The breastfeeding quality is measured using the BBAT tool,<sup>a</sup> which assigns scores (0–2) for four key breastfeeding components: position, attachment, sucking, and swallowing, yielding a total score of 0–8, with a higher score indicating better breastfeeding performance. We have defined the following categories: Poor (0-2), Moderate (3-6), Good (7-8).<br>**Measurement:** BBAT will be assessed by trained and standardised female IOA assessors, masked to group allocation. |

**Table 2.** Secondary Outcome Measures.

| Name | Definition |
| --- | --- |
| **Outcome 2.1:** Exclusive Breastfeeding at day 28 of birth. | **Description:** As per WHO, exclusive breastfeeding means that the infant receives only breast milk, with no other liquids or solids, not even water – with the exception of oral rehydration solution, or drops/syrups of essential medicines/ vitamins.<sup>a</sup><br>**Measurement:** This will be assessed via a 24-hour dietary recall, in a manner consistent with the Infant and Young Child Feeding indicators recommended by WHO and UNICEF. |
| **Outcome 2.2:** Breastfeeding Experience Scale Score measured on follow-up day 8. | **Description:** The Breastfeeding Experience Scale is a 19-point scale, which captures mothers' breastfeeding-related challenges (e.g., sore nipples and other breast-related problems, infant-related challenges to breastfeeding success, maternal anxiety, perceived breastfeeding success etc.). Each item is scored on an ordinal scale; higher scores indicate more negative breastfeeding experiences. <sup>a</sup><br>**Measurement:** It will be assessed by trained and standardised female IOA assessors, and the total score used as a continuous measure. |
| **Outcome 2.3:** Incidence of Possible Serious Bacterial Infection (PSBI) among infants on follow-up days (days 4, 8, 15 and 29). | **Description:** Possible serious bacterial infection commonly known as PSBI is defined as the presence of any of the following 7 clinical signs in the infants:<sup>a</sup><br>• not feeding well or not able to feed at all<br>• movement only when stimulated or no movement at all<br>• high body temperature (38 °C or above) |
|  | • low body temperature (< 35.5 °C)<br>• severe chest indrawing<br>• convulsions<br>• fast breathing (≥ 60 breaths per minute) in infants aged 0–6 days<br>**Measurement:** Clinical assessment by trained and standardised IOA team during follow-up visits as per IMNCI assessment protocol. A newborn with PSBI signs on any of the day 4, 8, 15 and 29 visits will be categorised as having PSBI during the follow-up period. |
| **Outcome 2.4:** Incidence of Maternal depression measured at 15 days after birth. | **Description:** The Edinburgh Postnatal Depression Scale (EPDS) consists of 10 short statements. Responses are scored 0, 1, 2 and 3 based on a Likert scale.<br>**Measurement:** The EPDS will be administered by trained and standardised female IOAs on the Day 15 follow-up visit. Mothers scoring above 11 (lower, more sensitive cut-off value) as per the EPDS scoring scheme will be considered as likely to be suffering from depression.<sup>a</sup> |
| **Outcome 2.5:** Mother-to-infant bonding scale score measured on day 29. | **Description:** The Mother-to-infant Bonding Scale (MIBS), consists of 8 different self-reported feelings of mothers towards their babies, and aims to assess how often mothers experience these feelings towards their babies. Each of the 8 items is scored on a 0-3 Likert scale, with lower scores indicating better attachment.<sup>a</sup><br>**Measurement:** The MIBS will be administered on follow-up day 29 visit by trained and standardised female IOAs, and the total score will be used as a continuous measure. |
| **Outcome 2.6:** Breastfeeding self-efficacy scale (short form) score measured on day 29. | **Description:** Maternal breastfeeding self-efficacy (BSES) measured by the BSES-Short Form consisting of 14 items scored on a 5-point Likert scale, with higher scores indicating greater confidence.<sup>a</sup><br>**Measurement:** The BSES-SF will be administered to mothers by trained and standardised female IOAs, and the total score will be used as a continuous measure. |

### Severe Adverse Events

Any severe adverse event (SAE) occurring after enrolment will be documented using standardised adverse event reporting forms. For this trial, newborn death or admission to a Level-2 or higher newborn care unit are classified as SAEs.

The SESS team will document SAEs that occur during facility stay, while the Independent IOA team will document SAEs identified during follow-up visits. All SAEs will be reported to the study coordinator/investigator immediately. The Principal Investigator will verify all SAEs involving newborn death or higher-level unit admission and notify the Data and Safety Monitoring Board (DSMB) within 24 hours.

All SAEs will be summarised and communicated to the ethics committee monthly. The DSMB will conduct a masked interim analysis when 50% of participants complete follow-up, assessing the safety of the trial by comparing the SAE incidence, severity and patterns between the two study groups. Given the low-risk nature of the study, no formal stopping rules have been specified.

### Participant Timeline

All eligible mother-newborn dyads will be enrolled during labour or within four hours after birth and randomised to the intervention or control group. Follow-up assessments will be conducted during the facility stay at 24 hours (Day 1) and 48 hours (Day 2), followed by scheduled visits on Days 4, 8, 15, and 29. These visits will include newborn and maternal assessments as specified in Table 3. Participant follow-up will conclude at Day 29.

**Table 3.** SPIRIT 2025 Diagram of the Schedule of Enrolment, Interventions, and Assessments.

|  | Enrolment |  | Post Randomization |  |  |  |  | Close-out |
| --- | --- | --- | --- | --- | --- | --- | --- | --- |
| Timepoint | Labour | <4hr | D1 | D2 | D4 | D8 | D15 | D29 |
| **Enrolment** |  |  |  |  |  |  |  |  |
| Verbal Consent & Registration | X |  |  |  |  |  |  |  |
| Eligibility Screening |  | X |  |  |  |  |  |  |
| Informed Written Consent |  | X |  |  |  |  |  |  |
| Randomization & Enrolment |  | X |  |  |  |  |  |  |
| **Intervention or Comparator** |  |  |  |  |  |  |  |  |
| **Intervention group:** Prolonged KMC (≥8 hours daily SSC with exclusive breastfeeding) with breastfeeding in the KMC position, supported during the first 48 hours by the KMC team. |  |  | X | ----- | ----- | ----- | ----- | → |
| **Both groups:** Common Minimum Package, including initiation of breastfeeding through uninterrupted SSC in the first hour, and counselling on essential newborn care. |  |  | X | X |  |  |  |  |
| **Outcome Assessment** |  |  |  |  |  |  |  |  |
| A. Newborn weight |  |  | X | X | X | X | X | X |
| B. PSBI |  |  |  |  | X | X | X | X |
| C. Anthropometry Measurement (length, head circumference, MUAC) |  |  | X |  |  |  |  | X |
| D. Breastfeeding Observation (BBAT) |  |  |  |  |  | X |  |  |
| E. Feeding Module |  |  | X | X | X | X | X | X |
| F. Newborn Care Practices |  |  | X | X | X |  |  | X |
| G. Maternal history |  |  |  |  | X |  |  |  |
| H. Breastfeeding Experience |  |  |  |  |  | X |  |  |
| I. Maternal Depression |  |  |  |  |  |  | X |  |
| J. Mother-infant bonding |  |  |  |  |  |  |  | X |
| K. Breastfeeding Self-Efficacy |  |  |  |  |  |  |  | X |
| L. Socio Economic Status |  |  |  |  |  | X |  |  |

### Sample Size

The sample size was calculated based on the primary outcome of percentage weight change over the first 48 hours of life, and assessed for adequacy against the remaining two primary outcomes. Using pilot data from a previously conducted community-based study, we assumed that normal birthweight infants lose a mean of 6% and standard deviation (SD) 3.6%, of their birthweight within the first 2 days of life. To detect a 25% relative reduction in early weight loss with 95% confidence, 90% power, and a design effect of 2.0, we would require 244 infants in each group, i.e., ∼122 infants per group in each facility.

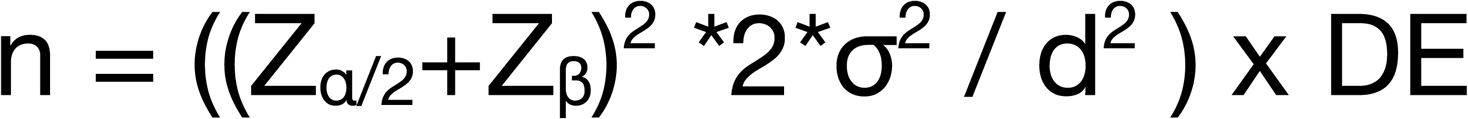

where Z_α/2_ is the critical value of the Normal distribution at α/2 (e.g. for a confidence level of 95%, α is 0.05 and the critical value is 1.96), Z_β_ is the critical value of the Normal distribution at β (e.g. for a power of 90%, β is 0.1 and the critical value is 1.28), σ^2^ is the population variance, and d is the difference between the two groups (25% of 6% = 1.5%). DE is the design effect due to enrolment at multiple sites, assumed to be 2.0. Assuming a loss to follow-up of 5%, we will need to enrol 129 infants per group in each facility, i.e. 258 infants in both groups per facility, giving a total sample size of 516 normal birthweight infants.

This sample size also provides ≥80% power to detect:

- a 20% relative improvement in weight gain velocity during the newborn period (at 95% confidence and 80% power, assuming control infants gain a mean of 25% of birthweight [SD 10%] over the first 28 days).
- a 50% reduction in the proportion of mother-baby dyads with a poor to moderate BBAT score of < 7 in the intervention arm as compared to control arm at 95% confidence level and 80% power (assuming 30% of dyads in the control group have a BBAT score of < 7).

## METHODS: ASSIGNMENT OF INTERVENTIONS

### Screening and Consenting

Pregnant women admitted to the study hospitals for delivery will be registered for post-delivery screening for study eligibility if they reside in the catchment area, are willing to stay in the hospital for the government-recommended 48 hours post-delivery, intend to breastfeed for the first 28 days, and provide verbal consent for the screening.

After delivery, all registered women will be supported in the initiation of breastfeeding through uninterrupted SSC within the first hour. Thereafter, a trained hospital nurse will measure the birth weight of their newborn using a calibrated Seca 334 weighing scale in the labour room, under the observation of study nurses. Additional screening information will be obtained from hospital records and verified with treating clinicians. A study nurse, in consultation with the on-duty paediatrician, will conduct a physical examination to identify congenital anomalies prior to confirming eligibility for enrolment.

For eligible mother-infant dyads, written informed consent will be obtained within 4 hours of birth. The study team will assess the mother’s readiness in consultation with the treating physician or nurses, and will ensure that the consenting process supports maternal comfort and comprehension and allows adequate time for questions.

### Randomisation, Allocation and Recruitment

Randomisation for this open-label individually randomised trial will be stratified by site. Allocation sequences for each site will be generated using a randomisation scheme with random permuted blocks of variable sizes (2,4,6) through a Python program. The file containing the allocation sequence for each facility will be electronically securely stored in the secure REDCap application backend database. During the enrolment process after securing informed written consent, the REDCap application will automatically assign the infant to the study group corresponding to the next sequence number and record the assigned group in the database, without making it visible in the case report forms. The Screening, Enrolment, and Systems Strengthening (SESS) team, consisting of trained nurses, will conduct the screening and enrolment.

As an open-label trial, the SESS team, KMC intervention support team, and participating mothers will be aware of group assignment, given the challenge of masking KMC as an intervention from those who are practising or supporting it. To minimise contamination, intervention and control mothers will be kept in separate but similar-looking and similarly equipped wards during their hospital stay. The SESS team will support the standardised provision of a common minimum package of care to mother-baby dyads in both study groups, to ensure basic quality of care even for mother-newborn dyads in the control group. Clear delineation of responsibilities among the participant-facing teams has been done to minimise bias (Table 4). All outcomes will be assessed through standardised procedures by an Independent Outcome Assessment (IOA) team masked to group allocation. Quality control measures will be adopted to ensure that the standard operating procedures are being duly followed and any deviation from the protocol will be documented.

**Table 4.** Composition and responsibilities of study teams.

| Screening, Enrolment and Systems Strengthening Team (SESS) | Intervention Support Team (KMC team) | Independent Outcome Assessment Team (IOA) |
| --- | --- | --- |
| Team consists of trained nurses supervised by the site coordinator in each facility.<br>Participant Identification and registration.<br>Obtaining verbal consent for screening of all delivered neonates at the study facility according to inclusion and exclusion criteria.<br>Ensuring SSC and breastfeeding initiation in the labour room for all registered mothers, followed by accurate measurement of birth weight.<br>Communicate with the treating nurse /physician to validate clinical findings, and complete the screening case report form.<br>Obtain written consent for enrolment and fill the required case report form, initiate randomisation in REDCap. Share the assigned study arm with the | Team consists of lay health workers trained in providing support for KMC, supervised by the site coordinator in each facility, and mentored by a technical specialist in KMC.<br>Provide counselling and hands-on support to mothers during their hospital stay for practising KMC and newborn-led latching for breastfeeding in the KMC position without any interference.<br>Maintain logs for SSC and breastfeeding.<br>Conduct telephonic reminders with mothers/ family members on the day after discharge, to encourage adherence to prolonged KMC (ideally $\geq 20$ hours, minimum $\geq 8$ hours daily). | Team consists of a male-female pair of trained workers to facilitate anthropometric assessment as well as sensitive tools (breastfeeding assessment, postnatal depression) that can only be administered by female workers. The IOA team is directly supervised for quality assurance by the project lead.<br>Conduct follow-up assessment for all enrolled mother-baby dyads on days 1, 2, 4, 8, 15 & 29, as per the schedule (see Table 3), and complete the required case report forms.<br>In case of any serious adverse event identified during follow-up visit, fill the required case report |
| site coordinator for requisite follow-up of intervention group mothers by the Intervention support team.<br>• Deliver common minimum counselling package in a standardised manner for both the study groups<br>• In case of any serious adverse event prior to discharge, fill the required case report form for further investigation. |  | form for further investigation. |

## METHODS: DATA COLLECTION, MANAGEMENT AND ANALYSIS

### Data Collection

All data collection will be done via Android-based tablet devices on the REDCap platform. The schedule of data collection is presented in Table 3. Data collection timepoints include enrolment, 24 hours (±6 hours) after birth (day 1), 48 hours (±6 hours) after birth (day 2), and days 4 (i.e. after 3 completed days of life), 8, 15 and 29. At the time of enrolment, the birth weight will be measured by facility nurses in the presence of the SESS team, and relevant screening information recorded by the SESS team. All case report forms for outcome assessment will be masked to the allocation. The IOA team with paired male and female assessors will conduct outcome assessment for all mother-baby dyads as per the data collection schedule through physical visits to the facility or at home after discharge. Anthropometric measurements will be conducted jointly by the two assessors, culturally sensitive assessments (e.g. breastfeeding scales, depression, etc.) conducted by female assessors, and neutral assessments (e.g., PSBI, socioeconomic status) conducted by the male assessors. The administration of the feeding and newborn care module (including SSC episodes) to mothers face-to-face during days 1 and 2, i.e. hospital stay, and administration of the newborn care module telephonically on day 4 will be conducted by the SESS team as an exception to preserve masking of the IOA team to the study group.

#### Participant retention

To maximise retention and complete follow-up, calls will be made to participants one day prior to each scheduled visit. Flexible visit windows (within 2 days of the scheduled follow-up day for home visits) will be allowed to accommodate participant availability. For participants who discontinue the intervention, outcome data will continue to be collected at all scheduled time points unless consent is withdrawn. Reasons for discontinuation or loss to follow-up will be documented.

### Training, Standardisation & Quality Assurance

All study team members will be trained and certified on Good Clinical Practice (GCP) guidelines. Prior to study initiation, a comprehensive site readiness assessment will be undertaken to standardise clinical practices, counselling procedures, measurements and documentation.

#### KMC Team

Team members will receive a 2-week hands-on training by a technical specialist on KMC – including safe positioning and breastfeeding in KMC, use of binders, counselling and supporting mothers, and maintaining a log of SSC sessions. The technical specialist will be available for on-call support and conduct weekly reviews with the KMC team to discuss cases and resolve queries and challenges. One-day refresher training will be conducted each month.

#### SESS Team

Initial 5-day training will focus on rapport-building and communication with mothers and caregivers, consenting procedures, eligibility assessment, standardised newborn weighing with calibration, and accurate completion of study forms. Subsequently, site coordinators will provide handholding support for 2 weeks. The SESS team will be supervised by site coordinators, who will conduct process observations for each team member every 14 days.

#### IOA Team

The IOA team will receive a 2-week training and standardisation, covering anthropometric measurements, each of the scales and other questionnaires, as well as accurate completion of study forms. Subsequently, they will be provided close handholding support on the field by the project lead over 2 weeks to ensure confidence in administration of each of the scales. The project lead will directly supervise the IOA team through scheduled and random visits. As part of quality assurance, a random 10% sample of study visits will be subjected to detailed spot checks. These reviews will assess protocol adherence, accuracy of data collection, and standardization of procedure implementation to ensure the reliability and integrity of study conduct across all sites.

### Data Management

Robust data management practices will be implemented to ensure data accuracy, integrity, and confidentiality. Data collected on the REDCap platform will be linked to a centralised, secure, cloud-based database accessible only to authorised personnel. The forms will have inbuilt checks for missingness, range, skips, etc. Data will be reviewed for range and consistency checks with weekly audits. Regular backups of electronic data will be maintained in separate secure locations, while physical records, such as consent forms, will be stored in locked cabinets with restricted access. All participant data will be assigned unique study identification codes and anonymised prior to any data sharing.

### Statistical Methods

#### General Principles

All statistical analyses will be performed using Python (Python Software Foundation, https://www.python.org/), with R software (R Foundation for Statistical Computing, https://www.r-project.org/) or STATA (StataCorp, College Station, Texas), as needed. The primary analysis will be conducted on an Intention-to-Treat (ITT) basis, including all randomised mother-infant dyads in the groups to which they were originally assigned, regardless of adherence to the intervention or protocol deviations. Statistical significance will be defined as a two-sided p-value ≤0.05.

This trial specifies three co-primary outcomes that collectively characterise the multidimensional process of early newborn adaptation: early metabolic stabilisation (percentage early weight loss, proximal outcome), feeding quality (BBAT score, intermediate outcome), and overall growth trajectory (weight gain velocity, distal outcome). These outcomes represent complementary domains of a single underlying construct, “successful neonatal transition”, rather than independent endpoints. Accordingly, no formal multiplicity adjustment will be applied to the primary analysis, consistent with recommendations for trials with conceptually related outcomes.^33,34^ However, we will interpret results holistically. Success will be adjudicated based on the totality of evidence, prioritising the directional consistency and clinical magnitude of effects across these three domains rather than a binary assessment of a single p-value. Secondary outcomes are considered exploratory; their results will be interpreted descriptively to provide context to the primary findings, with 95% confidence intervals reported without adjustment for multiplicity.

### Analysis Populations

- **Intention-to-Treat (ITT) Population:** All randomised participants. This will serve as the primary analysis population to estimate the efficacy of the intervention under real-world conditions.
- **Per-Protocol (PP) Population:** A secondary analysis will be performed restricting the intervention group to "compliers", i.e., participants with high fidelity to the protocol. Compliance is defined as achieving **≥**8 hours of daily skin-to-skin contact with exclusive breastfeeding performed in the KMC position in more than 80% of episodes during the hospital stay (first 48 hours).

#### Descriptive Statistics

Baseline demographic and clinical characteristics will be summarised by treatment group. Continuous variables will be presented as means with standard deviations (SD) or medians with interquartile ranges (IQR) based on the normality of distribution (assessed via the Shapiro-Wilk test). Categorical variables will be presented as frequencies and percentages. Baseline balance will be assessed clinically, and any significant imbalances will be considered for inclusion as covariates in adjusted models.

### Analysis of Primary Outcomes

- **Percentage Weight Loss & Weight Gain Velocity (Continuous):** The mean percentage weight loss at 48 hours and weight gain velocity during the newborn period will be compared between the intervention and control groups using Linear Mixed-Effects Models (LMM). The model will include the treatment group as a fixed effect and the study site as a random effect to account for stratification. To increase the precision of the estimated treatment effect, the LMMs will be adjusted for prespecified covariates that are strong biological predictors of the outcome: maternal parity (primiparous vs. multiparous), infant sex, and baseline birthweight. If residuals are non-normally distributed, 95% Confidence Intervals (CI) will be derived using Bias-Corrected and Accelerated Bootstrapping (1,000 replications).
- **· Quality of Breastfeeding (Binary):**

- Primary evaluation (Binary) – As per the study hypothesis, BBAT scores will be dichotomised into "Good" (≥7) vs. "Moderate-to-Poor" (<7). The proportion of dyads with moderate-to-poor scores will be compared using Mixed-Effects Logistic Regression, with study site included as a random intercept and adjustment for parity, infant sex and baseline birthweight. Adjusted odds Ratios (aOR) and 95% CIs will be reported.
- Sensitivity analysis (Ordinal scale) – To leverage the full information of the 0–8 scale, the raw BBAT score will be analysed using Mixed-Effects Ordinal Logistic Regression (Proportional Odds Model). This analysis will assess the intervention’s effect on achieving higher quality scores across the entire scale.

### Analysis of Secondary Outcomes

- **Binary Outcomes (e.g., Exclusive Breastfeeding, Maternal Depression, PSBI):** Analysed using mixed-effects logistic regression or Chi-square tests where modelling is not feasible due to low event rates.
- **Continuous Outcomes (e.g., Bonding Scores, Self-Efficacy):** Analysed using linear mixed-effects models or independent t-tests (with robust standard errors) as appropriate.

#### Handling of Missing Data

Missingness patterns will be examined. If missing data for primary outcomes exceeds 5%, Multiple Imputation by Chained Equations (MICE) will be employed, assuming data are Missing At Random (MAR). Sensitivity analyses will be conducted using Complete Case Analysis to check the robustness of the results.

## METHODS: MONITORING & OVERSIGHT

### Steering Committee

This committee will include all Principal Investigators and Co-Investigators from participating sites. It will oversee the finalisation of standard operating procedures, case report forms, conduct of the study, including all study procedures to ensure smooth trial execution, and participate in the analysis and dissemination of results.

### Trial Coordination Centre

This unit will be established at the Community Empowerment Lab, and consist of the project lead, KMC technical specialist and data management team. All data will be centrally managed, and an enrolment progress dashboard along with quality metrics with de-identified information will be reviewed during Steering Committee meetings held on a monthly basis. The Trial Coordination Centre will provide direct oversight for all study operations, manage study data, and ensure high quality research execution as per protocol.

### ICMR Scientific Review Committee

A Scientific Review Committee at ICMR will provide scientific and ethical oversight and review the progress of the study. The program manager at ICMR may conduct site monitoring visits as required.

### Data Safety Monitoring Board (DSMB)

An independent DSMB consisting of 3 members with expertise in neonatology, biostatistics, and clinical trial methodology will monitor trial integrity, protocol adherence, and recruitment progress. The DSMB will review SAEs and provide recommendations to mitigate risks.

## ETHICS

### Research Ethics Approval

The study protocol has been approved by the Institutional Ethics Committees of the Community Empowerment Lab (CEL/RES/202507/001), which will provide direct oversight to the conduct of the study in two study sites – District Women’s Hospitals in Lucknow and Barabanki, and King George’s Medical University (130th ECM IIA/P20), which will provide direct oversight to the conduct of the study in KGMU. Protocol amendments, if at all required, will be submitted for review and approval to the relevant ethics committees prior to implementation. Approved amendments will be communicated to all study sites, updated in the trial registries (CTRI and ISRCTN), and reported to the study sponsor. The study will follow Good Clinical Practice guidelines and ethical principles outlined in the Declaration of Helsinki. The safety of enrolled infants will be closely monitored throughout the trial through structured follow-up and adverse event reporting.

### Informed Consent

Verbal consent will be sought from mothers prior to birth to perform screening procedures. After screening, written informed consent will be obtained from the mothers of newborns who meet the eligibility criteria for enrolment. The consenting process will be conducted by trained study staff in the participant’s preferred local language. For illiterate participants, consent will be documented by a thumbprint in the presence of an impartial witness. Mothers will be made aware of the study’s purpose, procedures, potential risks and benefits, and the voluntary nature of participation, as well as their right to withdraw at any time without repercussions.

### Confidentiality & Data Protection

Confidentiality of participants and data protection will be strictly upheld, with data anonymised and securely stored. Any adverse events or unintended effects of the intervention will be carefully monitored, documented, and addressed promptly. Only anonymised data will be made available for analysis, sharing and dissemination.

### Ancillary and Post-trial Care

The study involves minimal risk, as it evaluates KMC, a safe and established intervention in routine newborn care. Given the minimal-risk nature of the study, no specific compensation or insurance is planned. In the unlikely event of any study-related adverse event, appropriate medical care will be provided as standard facility norms. All participants will continue to receive standard newborn care during the study.

## DISCUSSION

The transition from intrauterine to extrauterine life represents a critical physiological window where neonates must rapidly adapt to new thermal, respiratory, and metabolic demands.^1,3,10^ While Kangaroo Mother Care (KMC) is the established gold standard for supporting this transition in preterm and low birthweight infants with multiple proven benefits beyond survival alone,^19^ its potential benefits for healthy, normal birthweight newborns remain significantly underexplored. This protocol describes the first multicentre randomised controlled trial to systematically evaluate the efficacy of prolonged KMC (combining skin-to-skin contact and exclusive breastfeeding in KMC position) during the critical first 72 hours of life, specifically for normal birthweight infants.

A key strength of this trial is its pragmatic, multicentre design within public health facilities, which enhances the external validity and scalability of the findings. The first 72 hours is the peak period for early weight loss, coinciding with early metabolic adaptation and the establishment of lactogenesis II, when maternal-newborn SSC may have the greatest physiological and behavioural influence.^5,10,11^ The recommended daily duration of SSC in the study is aligned with the WHO recommendations for preterm and low birthweight infants to ensure consistency. The primary outcomes are carefully chosen to capture core domains relevant to plausible mechanistic pathways for benefit: percentage weight change at 48 hours as an indicator of metabolic stabilisation; weight gain velocity over the first 28 days as a measure of early growth trajectory; and the Bristol Breastfeeding Assessment Tool score at seven completed days as a validated marker of breastfeeding quality. In combination with the secondary outcomes, the trial will generate comprehensive evidence on the effects of prolonged KMC in the mother-newborn dyad.

We acknowledge certain limitations inherent to the study design. The open-label nature of the trial is unavoidable given the visible nature of the intervention; however, the protocol mitigates bias through several measures, including an independent outcome assessment team masked to group allocation and ensuring a basic minimum standard of care for all enrolled newborns. Given the nature of the intervention, we will need to provision reasonable support through intervention workers to ensure intervention quality and reliable assessment of efficacy. The potential Hawthorne effect will be mitigated by utilising lay health workers for intervention support, and through equivalent visits by study nurses to both intervention and control group mothers during their hospital stay. Being the first study of its kind, we will exclude newborns born through caesarean section and complicated births to simplify the intervention, analysis and subsequent interpretation. We chose to apply birthweight as a measure for assessing eligibility and not gestational age or a combination of the two, due to the variable quality of gestational age information in our study setting.

This trial will provide the first rigorous evidence on whether sustained KMC in the immediate postnatal period benefits healthy, normal birthweight infants – a population that represents the majority of newborns yet remains largely absent from prior KMC research.

The current recommendation for normal birthweight infants involves uninterrupted skin-to-skin contact immediately after birth for about an hour until breastfeeding is initiated.^18^ Biological plausibility suggests that extending the duration of SSC to the first 72 hours and beyond may also benefit normal birthweight infants, such as improved thermal and cardiorespiratory stabilisation, breastfeeding, growth, mother-infant bonding, and neurodevelopment, as observed in LBW infants. By evaluating the safety, efficacy, and feasibility of KMC for normal birthweight infants, the study could have significant implications for clinical practice and guidelines.

If successful, our findings could challenge the prevailing paradigm that KMC is solely a risk-reducing intervention meant for vulnerable small babies, repositioning it as a salutogenic essential standard of care for *all* newborns.^35^ The results may also guide the design of future research on KMC dose, applicability following caesarean birth, longer-term developmental effects, and contribute to a broader reconsideration and consolidation of recommendations for early postnatal care for mothers and normal birthweight infants across diverse settings.

## Supporting information

SPIRIT CHECKLIST

## Data Availability

Since this is a study protocol, this is not applicable.

## ACKNOWLEDGEMENTS

We extend our sincere thanks to Dr Taruna Madan, Head, and Dr Gunjan Kumar, Scientist D, Development Research Division, Indian Council of Medical Research, for their administrative oversight and support throughout the study. We also thank the Ministry of Health and Family Welfare, Government of Uttar Pradesh, and the National Health Mission for their collaboration and facilitation of study implementation within public health facilities. We thank the Scientific Committee of the Indian Council of Medical Research (ICMR) – Dr Shrikant Tripathi (Director Medical Research, Dr. D. Y. Patil Medical College, Hospital & Research Centre, Pune), Dr Suman Kanungo (Director, ICMR - Regional Medical Research Centre, North East, Dibrugarh), Dr Devendra Mishra (Professor, Department of Pediatrics, Maulana Azad Medical College, Delhi), and Dr Aparna Mukherjee (Scientist F and Head, Clinical Studies and Trials Unit, Development Research Division, Indian Council of Medical Research), for their technical inputs and guidance. We thank the members of the Data Safety Monitoring Board (DSMB) – Dr Jai Vir Singh (Chairperson, DSMB; Former Dean and Head of Department, Community Medicine & Public Health, King George’s Medical University, Lucknow), Dr Shambhavi Mishra (Assistant Professor, Department of Statistics, Lucknow University), and Dr Amit Kumar Gond (Paediatrician and Neonatologist, Department of Paediatrics, Sanjay Gandhi Hospital, Amethi) for their independent oversight and guidance.

## AUTHOR CONTRIBUTIONS

- **Conceptualisation:** Aarti Kumar, Vishwajeet Kumar
- **Methodology:** Aarti Kumar, Vishwajeet Kumar, Rashmi Kumar, Ashok Kumar, Gary Darmstadt
- **Writing – original draft:** Aarti Kumar, Malvika Mishra, Madhuri Tiwari
- **Writing – review & editing:** Rupak Mukhopadhyay, Vinay Pratap Singh, Anushka Srivastava, Mala Kumar, Anjoo Agarwal, Shakal Narayan Singh, Shalini Tripathi, Mohd. Salman Khan, Pradeep Kumar

